# A System Dynamics Model for Effects of Workplace Violence and Clinician Burnout on Agitation Management in the Emergency Department

**DOI:** 10.1101/2021.06.16.21258632

**Authors:** Ambrose H. Wong, Nasim S. Sabounchi, Hannah R. Roncallo, Jessica M. Ray, Rebekah Heckmann

**Affiliations:** Department of Emergency Medicine, Yale School of Medicine, New Haven, CT, USA; Graduate School of Public Health and Health Policy, City University of New York, New York, NY, USA; Department of Emergency Services, Yale New-Haven Hospital, New Haven, CT, USA

## Abstract

**Background:** Over 1.7 million episodes of agitation occur annually across the United States in emergency departments (EDs), some of which lead to workplace assaults on clinicians and require invasive methods like physical restraints to maintain staff and patient safety. Recent studies demonstrated that experiences of workplace violence lead to symptoms of burnout, which may impact future decisions regarding use of physical restraints on agitated patients. To capture the dynamic interactions between clinicians and agitated patients under their care, we applied qualitative system dynamics methods to develop a model that describes causal feedback mechanisms of clinician burnout and the use of physical restraints to manage agitation.

**Methods:** We convened an interprofessional panel of clinician stakeholders and agitation experts for a series of model building sessions to develop the current model. The panel derived the final version of our model over ten sessions of iterative refinement and modification, each lasting approximately three to four hours. We incorporated findings from prior studies on agitation and burnout as a result of workplace violence, identifying interpersonal and psychological factors likely to influence our outcomes of interest to form the basis of our model.

**Results:** The final model resulted in five main sets of feedback loops that describe key narratives regarding the relationship between clinician burnout and agitated patients becoming physically restrained: (1) use of restraints decreases agitation and risk of assault, leading to increased perceptions of safety and decreasing use of restraints in a balancing feedback loop which stabilizes the system; (2) clinician stress leads to a perception of decreased safety and lower threshold to restrain, causing more stress in a negatively reinforcing loop; (3) clinician burnout leads to a decreased perception of colleague support which leads to more burnout in a negatively reinforcing loop; (4) clinician burnout leads to negative perceptions of patient intent during agitation, thus lowering threshold to restrain and leading to higher task load, more likelihood of workplace assaults, and higher burnout in a negatively reinforcing loop; and (5) mutual trust between clinicians causes increased perceptions of safety and improved team control, leading to decreased clinician stress and further increased mutual trust in a positively reinforcing loop.

**Conclusions:** Our system dynamics approach led to the development of a robust qualitative model that illustrates a number of important feedback cycles that underly the relationships between clinician experiences of workplace violence, stress and burnout, and impact on decisions to physically restrain agitated patients. This work identifies potential opportunities at multiple targets to break negatively reinforcing cycles and support positive influences on safety for both clinicians and patients in the face of physical danger.

## BACKGROUND

Behavioral health conditions increasingly present to the emergency department (ED). In the United States, there has been an estimated 53% increase in number of mental health-related ED visits over the past decade, while overall visits only rose by 8.6%.^1^ Agitation, defined as excessive psychomotor activity leading to aggressive and violent behavior,^2^ is often part of these patient encounters, with 1.7 million episodes treated annually in EDs^3^ nationwide. Once agitation has occurred, clinicians are required to rapidly diagnose potential causes and intervene to minimize harm. However, treatment of these agitation episodes poses significant threats to safety for both patients themselves and ED clinicians caring for them. As such, physical restraints may be indicated and necessary if imminent danger for patients and staff are present. Although physical restraints are commonly used in the ED, physical trauma, significant respiratory depression, and asphyxiation leading to cardiac arrest can develop from restraint use.^4-6^ Concurrently, healthcare workers are increasingly at risk for workplace violence (WPV) while caring for agitated patients, with the ED identified as one of the highest risk environments.^7^ In a survey-based study, 78% of emergency physicians reported being targets of verbal and physical assaults at the workplace in the previous 12 months^8^ while >80% of Emergency Nurses Association members reported being victims of physical and verbal abuse while on shift.^9^ Studies have demonstrated missed workdays as high as 135 episodes per 10,000 workers each year from ED WPV incidents.^10^

Expert consensus panels have created separate recommendations regarding minimizing use of restraints during management of agitation^3^ and prevention of WPV.^11^ However, translation of these recommendations into pragmatic interventions that improve safety in an evidence-based manner has been limited by challenges at the bedside.^7^ Multiple factors during the interaction between clinicians and patients influence development of agitation and workplace violence events. Hence, implementing individual solutions (e.g. improving de-escalation techniques, increasing event reporting) in isolation may be impeded by time, resource, and logistical constraints in the busy and unpredictable environment of emergency care.^12,13^ In addition, our previous work has demonstrated that agitation management and WPV are complex, interlinked issues that require a comprehensive and systematic approach to help policymakers develop strategies that lead to meaningful change at the bedside.^14^ Most importantly, we found that WPV and agitation management need to be considered together as one and the same issue to balance patient safety with prevention of staff assaults for any potential interventions to be effective.^15^

Recent studies demonstrated strong associations between ED clinicians’ experiences of WPV and symptoms of burnout.^16,17^ ED clinicians are particularly affected due to increased treatment of mental health conditions in emergency care and growing systems-level challenges, such as overcrowding and boarding of admitted patients.^18,19^ As a result, increasing reports of burnout are appearing in the literature, ranging from 60 to 71% of respondents in survey-based studies with emergency physicians.^20,21^ Given that clinicians have reported both suboptimal patient care due to burnout^22^ and feelings of frustration and negative attitudes towards agitated patients,^23,24^ potential relationships between clinician burnout and agitation management deserve further investigation.

In this work, we applied qualitative system dynamics (SD) methods to develop a model that captures the dynamic interactions between ED clinicians and agitated patients, specifically focusing on how workplace violence affects decisions to use physical restraints during agitation care as mediated through symptoms of clinician burnout. SD modeling has been extensively used for healthcare and public health applications to study the dynamic behavior of healthcare issues in a complex system and provide a framework to develop insights into policies and potential interventions.^25^ It is a rigorous methodology that studies the dynamic behavior of a complex system by identifying its causal structure and feedback loops.^26^ It can be used to tackle the complexity of healthcare issues and test different policies to make better decisions for the future.^27^ SD models can integrate the key social, behavioral, and biological factors of interest into a single testable framework. They are therefore broad in scope and often include time delays, non-linearities, and behavioral feedback loops not included in typical statistical models.^28^ We hypothesize that SD modeling techniques can assimilate the growing body of knowledge regarding workplace assaults, clinician burnout, and use of physical restraints to determine potential strategies to optimize outcomes during agitation and workplace violence events in the ED.

## METHODS

### Study Design

In developing our qualitative SD model, we started by defining the scope and context of the problem to be studied and the goals of the modeling project. Our key initial planning discussions focused on balancing robust representation of the true complexities of agitation management with pragmatic development of an initial model that contains some acceptable limitations as a foundation for future work. Since our primary goal was to examine interactions between ED clinicians and patients during episodes of agitation, we bounded the clinical domain of our model by the context of an ED visit once a patient enters the physical space of an ED. We also selected the physician and nursing professions to represent the clinicians in this first iteration of our model since they often exert the strongest influence on key decisions around use of restraints.

To identify the overall goals of our SD modeling process, we next sought to define our problem in the form of a reference mode, indicated by a time-series graph that represents an abstraction of the most important variables that change over time in our model. We chose two key variables as primary outcomes of equal value, with rates of physical restraint representing patient safety and rates of clinician burnout representing staff safety. Although the literature has not clearly established temporal trends regarding rates of restraint use or clinician burnout in the ED, recent studies have demonstrated increasing numbers of ED visits for behavioral and mental health-related conditions over the past decade.^1,29,30^ Thus, we postulated that the number of agitation events is likely rising as well, leading to increasing rates of both restraint use and ED clinician burnout due to episodes of workplace violence. The corresponding graph for our reference modes is presented in **Figure 1**. Our feared trend within the reference mode would demonstrate increasing rates of restraint use and clinical burnout over time, while our hope is that potential interventions would flatten or even decrease rates of our outcomes over time.

**Figure 1.**
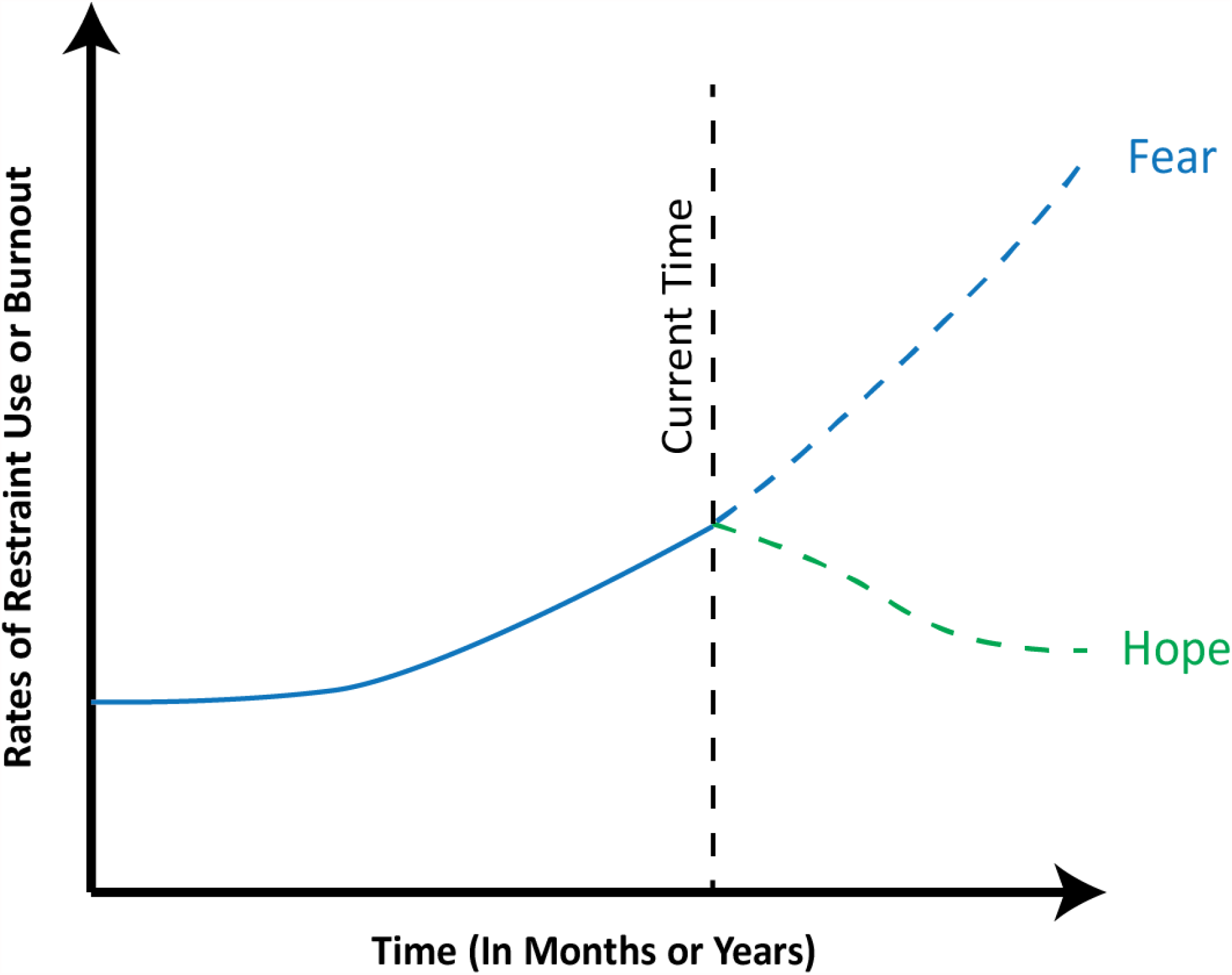
Reference Modes for our System Dynamics Model of Agitation Management. Key variables consist of rates of physical restraint use and rates of burnout. Feared trends indicate rising rates for both variables, while hope represents flattening or decreasing rates due to implementation of effective interventions over time.

In the next step, we created the qualitative causal loop diagram^31^ for our model, depicting the networks of causal factors and feedback loops relevant to the problem at hand. The resulting diagram of our model aims to summarize the mechanisms and alternative ideas for explaining the dynamics of interest within the context of the reference mode for the rates of restraint use and clinician burnout. This diagrammatic model serves as the first step of our overall SD modeling process and creates the foundation for mathematical simulations in future work.

Our model includes variable names and arrows with positive (+) and negative (–) signs consistent with standard SD practices. The arrows refer to our hypothesized causal relationships between individual variables over time. Positive signs indicate that two variables change in the same direction. Negative signs indicate that two variables change in opposite directions. A closed sequence of arrows (i.e., complete circles) form two kinds of feedback loops. The first type of feedback loops are *balancing loops* that serve to stabilize the system, bringing variables into steady states. A balancing loop has an odd number of negative links. The second type of feedback loops, *reinforcing loops*, can lead to exponential growth and build-ups in the system. A reinforcing loop has an even number of negative links in the model. These cycles can be positively reinforcing, or it can be negatively reinforcing, where a problem worsens over time, often at an increasing rate of speed.^32^

### Model Derivation and Data Analysis

To create a robust SD model for the management of agitation and clinician burnout, our team implemented a series of iterative modeling sessions with our interprofessional team of clinical stakeholders (nurses, physicians, hospital administrators) and researchers in agitation and workplace violence. Each structured session consisted of variable elicitation, derivation of behavior-over-time graphs, and illustration of variable connections via closed-loop diagrams. We incorporated two datasets from our previously published findings on systems approaches to agitation and workplace violence^14,15,33^ as well as current literature regarding burnout as a result of workplace violence,^9,17,34,35^ identifying interpersonal and psychological factors like mutual trust, perceptions of safety, and perceptions of team leadership that are likely to influence our outcomes of interest in measurable ways. We analyzed and interpreted data using principles of grounded theory in qualitative research.^36,37^ Three members of the research team (AHW, JMR, RH) started with a systematic, inductive approach to data analysis through an initial round of open coding to generate concepts “grounded” in participant views collected via field notes and still photographs of cognitive artifacts such as care timelines and diagrams. We then achieved consensus on major themes, model factors, and relationships through an iterative analytic process as more information was added after each modeling session using the constant comparative method.^38^ We collectively derived the final version of our model over ten sessions of iterative refinement and modification, each lasting approximately three to four hours. Our work received approval from our institutional review board as an exempt study.

## RESULTS

Our final SD model describes factors and relationships related to patient agitation, clinician burnout, and use of restraints that are known to contribute to the quality of patient care in the ED but that are infrequently mapped or explicitly described together. This model illustrates both the physical flow of patients through the ED and the actions of clinicians within the ED, in addition to demonstrating how factors influence outcomes for both groups. A simplified, high-level representation of the system is shown in **Figure 2** with four key sections of the model visually depicted with distinguishing colors. Subsequent **Figures 3 and 4** highlight a number of feedback loops that illustrate five key model narratives that describe groups of interdependent factors contributing to patient restraint and clinician burnout. We first discuss the overall model structure and its four key sections (A-D) and then focus on the five feedback loops as the main model narratives (1-5) in detail below. The model is depicted in a simplified version in Figures 2-4 so that relevant feedback loops and key insights can be more easily understood and described within this work. Please refer to the appendix for detailed versions of the full model that captures more detailed relationships and represents the synthesis of results from the modeling sessions.

**Figure 2.**
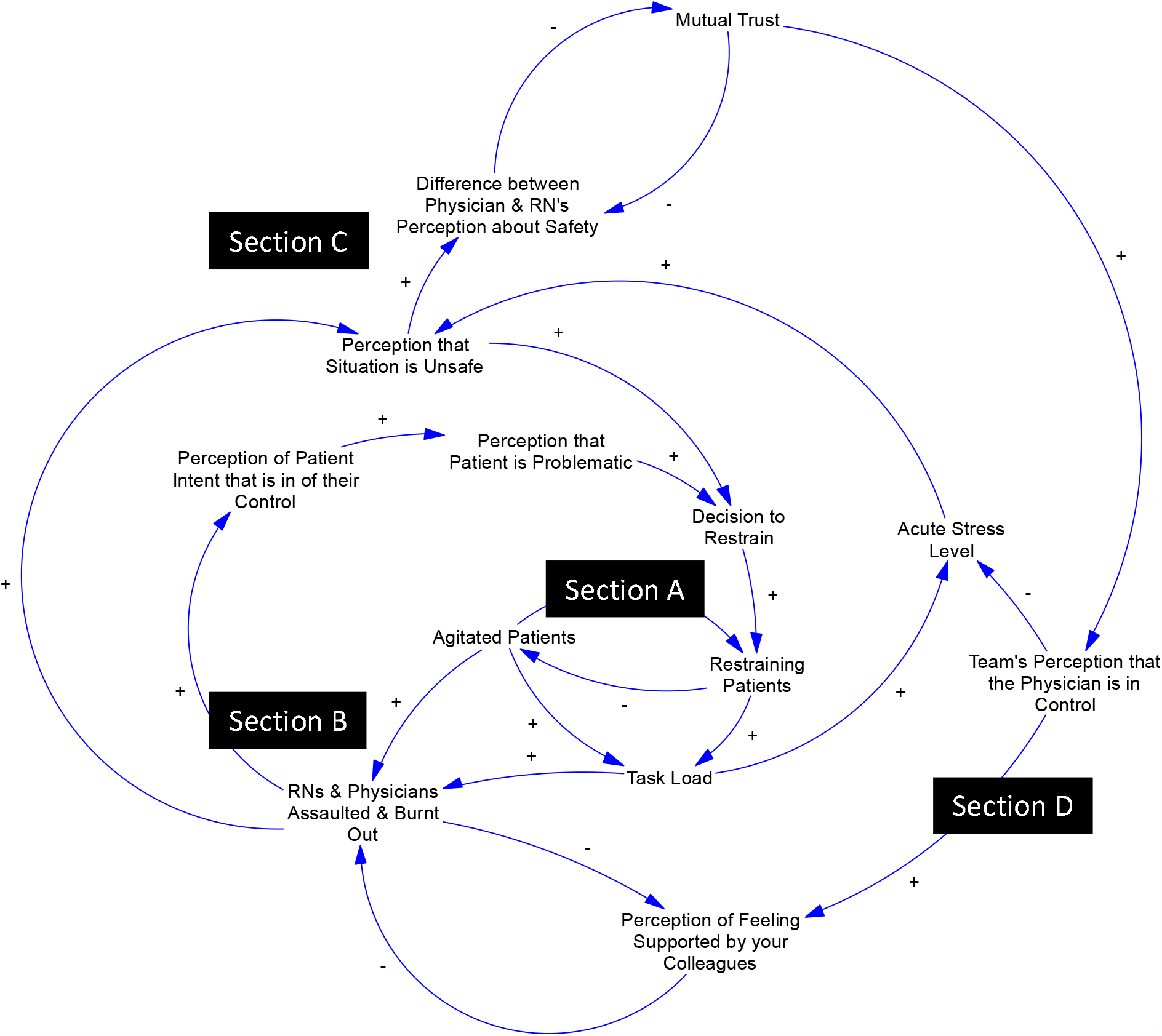
Qualitative System Dynamics Model for Agitation Management, Clinician Burnout, and Decisions for Physical Restraint Use. Sub-sections include A) Agitated Patients and Effects on Task Load; B) Clinicians Affected by Burnout and Assaults; C) Perceptions of Safety, Patients, and Development of Trust Between Members of the Team; and D) Perceptions of Control and Team Support.

**Figure 3.**
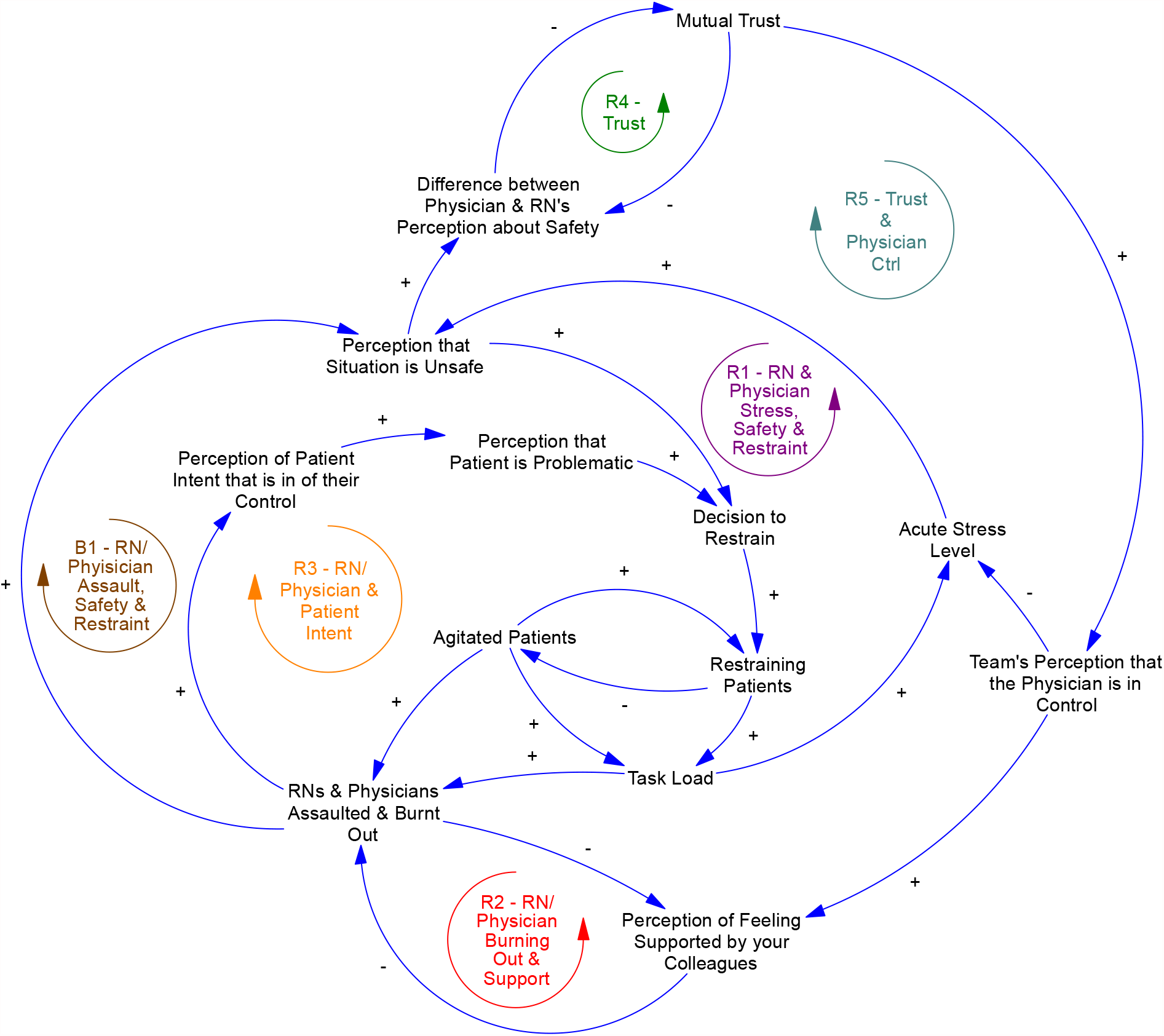
Highlighted Balancing and Reinforcing Loops. B1: Balancing Loop Involving Use of Restraints, Assaults, and Perceptions of Safety. R1: Negatively Reinforcing Loop Involving Clinician Stress, Safety, and Use of Restraints. R2: Negatively Reinforcing Loop Involving Clinician Burnout and Support. R3: Negatively Reinforcing Loop Involving Burnout & Perception of Patient Intent. R4: Positively Reinforcing Loop for Mutual Trust. R5: Positively Reinforcing Loop for Physician Control and Trust.

**Figure 4.**
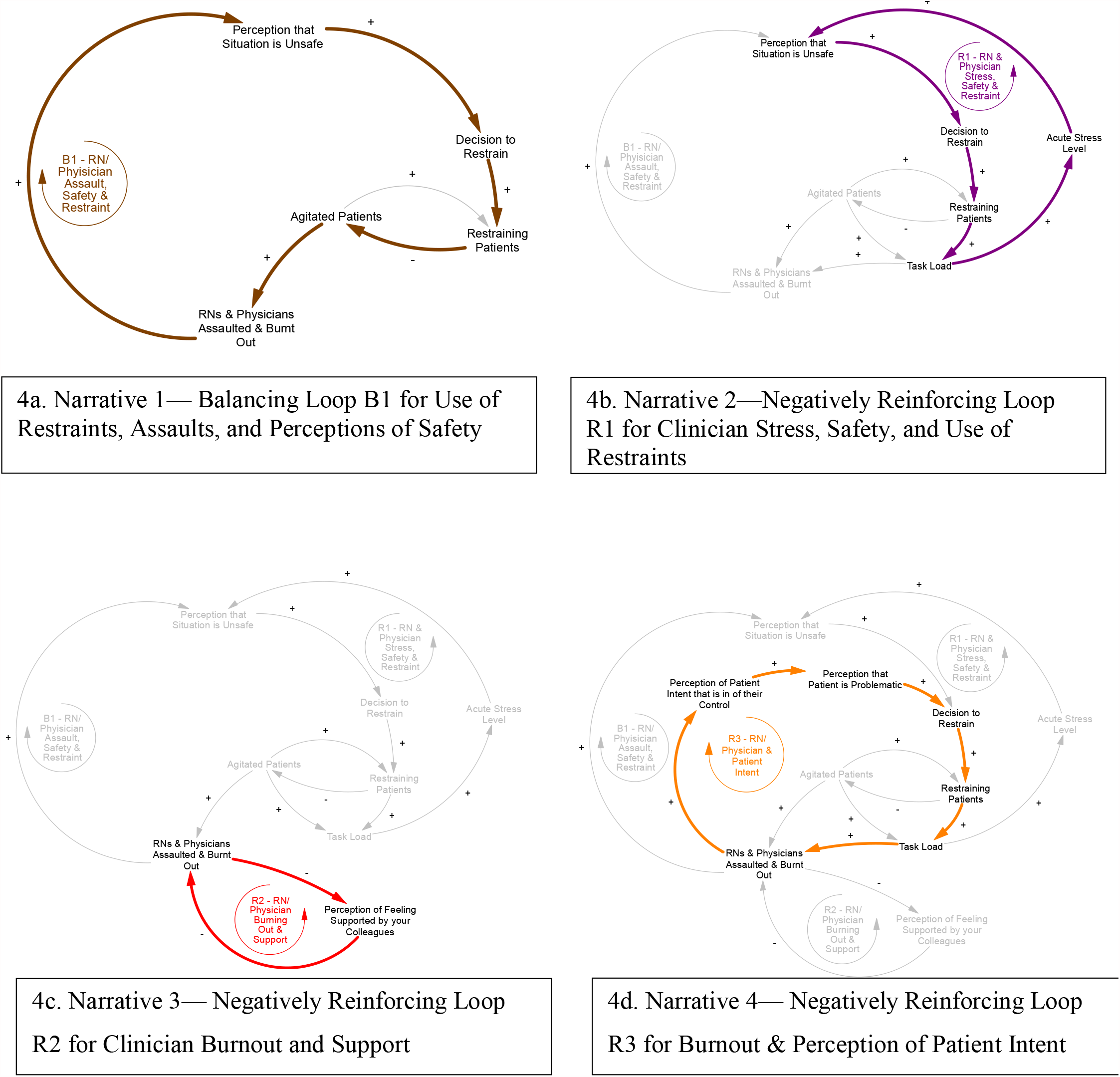
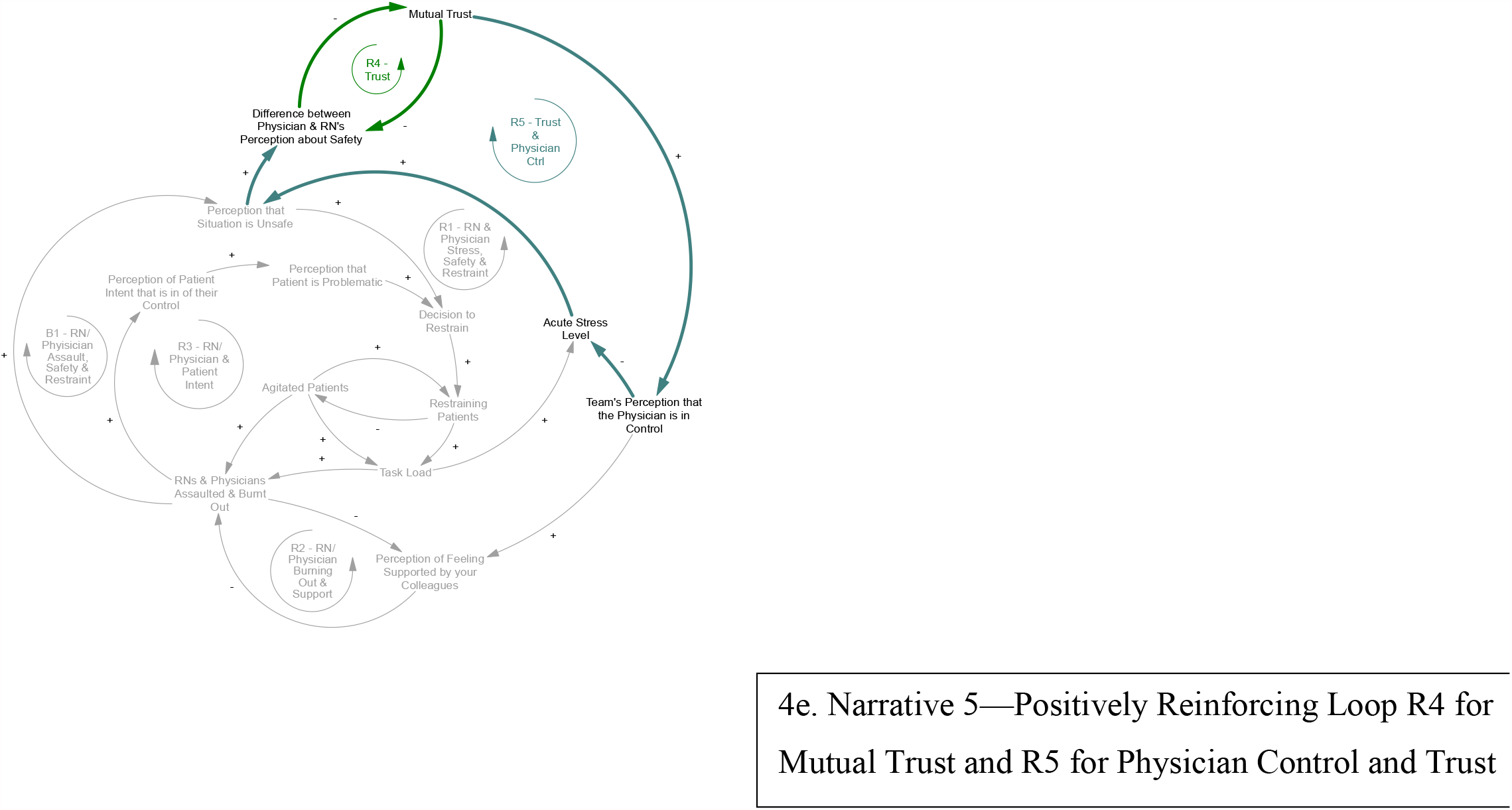
Individual Narratives (4a-4e).

### Model System and Structure (Figure 2)

**Section A** consists of the portion of the model addressing the movement of agitated patients through the ED and the corresponding effects on clinician task load. It illustrates how patients in an agitated state may become restrained. Importantly, both agitated patients and restraining patients add to clinician task load in different ways and at different time points of an encounter (e.g., bedside reassessment of patient condition and level of agitation). **Section B** depicts clinicians who may have varying experiences of being assaulted and/or be in varying stages of burnout. For simplification purposes, this cohort of clinicians is depicted as one group within the model. An increased number of either agitated patients or patients that are restrained may lead to increased likelihood of burnout or assault. We recognize that there may be a complex array of varying experiences and clinician profiles within that cohort (please refer to appendix for a more detailed depiction of this section of the model). We also recognize that clinicians may experience burnout for other reasons other than their interface with agitation events which will not be directly addressed within this model.

In **Section C**, the acute stress level of clinicians will contribute to a change in how each clinician views the safety of the current clinical encounter relative to the agitated patient. Similar perceptions of clinical safety between physicians and nurses will lead to increased mutual trust, while discrepancies in clinical perception of safety will lead to decreased mutual trust during and after a shift in the ED. **Section D** illustrates how each team will have varying perceptions regarding the physician’s control as the team leader. While different team structures are present at different hospitals and the concept of an ideal team structure may be a matter of debate, the physician with the most clinical experience is frequently in the role of team leader who is responsible for making final decisions regarding critically ill and agitated patients in many EDs throughout the country.^39,40^ In the model, the team’s overall perception that the physician is in control is shown to influence the acute stress level of clinicians and perceptions of feeling supported by colleagues.

### Model Narratives

One of the primary goals of model building was to identify balancing loops that stabilize the current system and contribute to the maintenance of clinical status quo, as well as reinforcing loops that magnify (positive or negative) effects on the system. The process of model building allowed us to identify five clinical narratives that illustrate how the presence of a given group of factors can lead to cycles of influence on each other in the form of one balancing loop and four reinforcing loops (**Figure 3**). We highlight these five main model narratives that describe our particular feedback loops of interest below. **Figure 4** displays pertinent factors and relationships for each narrative within individual subpanels (4a-4e) and sequentially adds more factors and relationships relevant to each subsequent narrative, building out the entire model in the fifth and final subpanel.

#### Narrative 1—Use of Restraints, Assaults, and Safety (Balancing Loop: Figure 4a)

This first narrative describes a balancing loop that serves to stabilize the system and likely reflects the instinctive and immediate reactions clinicians may have to protect themselves when faced with immediate threats to personal safety during agitation events. Loop B1 describes how a prior experience of assault is likely to increase a clinician’s perception that a given clinical agitation situation is unsafe, which, in turn, is likely to positively influence that clinician’s decision to place restraints during the clinical encounter. This increased likelihood to restrain then positively impacts the likelihood that an agitated patient becomes restrained, acutely decreasing the number of agitated patients as described in Section A of Figure 2. This, in turn, decreases the likelihood of clinician assault and burnout. Although this balancing loop through use of restraints to decrease assaults and increase perceptions of safety reflects a protective mechanism within the system to maintain restraint use and clinician burnout at steady states, the following narratives highlight negatively reinforcing loops that counteract the effects of this balancing loop to cause harm and explain why this balancing loop may not be sufficient.

#### Narrative 2—Clinician Stress, Safety, and Use of Restraints (Negatively Reinforcing Loop: Figure 4b)

In our model, clinicians develop increased acute stress as their task loads increase. These increased task loads decrease the perception of safety in the clinical environment. In turn, decreases in perception of safety lower the thresholds regarding decisions made by clinicians to restrain agitated patients, which then increase task load and acute stress. The notation R1 refers to the reinforcing loop that describe the relationship between clinician stress, perceptions of clinical safety, and use of restraints. Thus, increased clinician stress level leads to a perception of decreased safety and a lower threshold to restrain, causing more stress.

#### Narrative 3—Clinician Burnout and Support (Negatively Reinforcing Loop: Figure 4c)

The narrative embodied by the R2 reinforcing loop illustrate how both professions can become caught in negatively reinforcing cycles of burnout and decreased perception of colleague support. An increased number of clinicians who are assaulted and burnt out will contribute to a decreased perception of feeling supported by colleagues, which, in turn, will lead to an increased rate of burnout. Thus, clinician burnout leads to a decreased perception of colleague support which leads to more burnout.

#### Narrative 4—Burnout & Perception of Patient Intent (Negatively Reinforcing Loop: Figure 4d)

The narrative illustrated by the R3 reinforcing loop links clinician burnout to perceptions of patient intent. If clinicians have increased perceptions that agitated patients are “in control” of their aggressive behavior, referring to a perceived notion that the patient is displaying symptoms of agitation on purpose (e.g., to promote self-gain or maligned intent),^41^ perceptions that they are problematic could also rise as a result. The labeling of a patient as “problematic,” defined as an attribute of causing trouble projected onto a patient by a clinician, lowers a clinician’s threshold for decision to restrain, which, in turn, leads to an increased number of restrained patients. An increased number of restrained patients leads to increased clinician task load (e.g., during placement of restraints and subsequent reassessment and monitoring), which in turn contributes to an increased likelihood that nurses or physicians will be assaulted.^33^ As described in Section B of the model, clinicians can progress through different states of assault and burnout. An increased likelihood of assault ultimately leads to an increased number of clinicians who have been assaulted and who are burnt out. Finally, an increased number of clinicians with a history of assault and burnout will contribute positively to the perception that patients are in control of their agitated behavior.^23^ Thus, clinician burnout leads to negative perceptions of patient intent during agitation, lowering the threshold to restrain and leading to higher task load, more likelihood of workplace assaults, and higher burnout.

#### Narrative 5—Development of Trust and Control (Positively Reinforcing Loops: Figure 4e)

The narrative described by R4 refers to the positively reinforcing loop that illustrates the relationship between the different perceptions that nurses and physicians may have about clinical safety and the process of building trust. As the quantity of mutual trust builds between nurses and physicians, these clinicians are likely to have smaller differences in their perceptions of safety within a given clinical environment. A smaller discrepancy in perceptions of safety allows for the development of an increased amount of mutual trust, leading to a cycle of trust building. In addition, the narrative depicted by R5 illustrates that, in clinical environments where mutual trust between nurses and physicians increases, the team’s overall perception that the physician is effectively in control of the team rises as a result. This perception decreases the acute stress level of physicians and contributes to changes in physician perceptions about the safety of the clinical environment. As a result, the differences of safety perceptions between nurses and physicians decreases and feeds into mutual trust building as described by the R4 narrative. Thus, mutual trust between clinicians causes decreased discrepancy in perceptions of safety and increased perceived control of the team, leading to decreased clinician stress levels and further increased mutual trust.

## DISCUSSION

We developed a qualitative system dynamics model describing the complex interactions of ED workplace violence, clinician stress and burnout, mutual trust, and team orientation on physical restraint use in the management of patient agitation. Our model demonstrated the influence of individual clinician perceptions of work and safety, as well as team dynamics on decisions to restrain agitated patients. Recent surveys indicate that ED clinicians experience high rates of workplace violence and burnout.^8,42^ Direct threat of violence to staff can lead to heightened arousal and decreased productivity including changes in cognition and workload management.^43^ This work led to a dynamic framework for understanding and describing the conscious and unintended influences that prior experiences with workplace violence, burnout, team support, and mutual trust can have on management decisions during agitation.

While the decision to restrain an agitated patient occurs quickly, the factors influencing that decision may have developed over the course of a clinician’s shift, work week, year, or lifelong career. SD modeling allowed us to explore the change in clinicians’ management decisions over time as a factor of both near-term and long-term system changes. By uncovering key balancing and reinforcing loops within our model, we were able to identify points of interest for potential intervention that may otherwise have been hidden or buried within the complex interconnected factors of the system. One set of balancing loops reflect the instinctive reaction to restraint use that acts to stabilize the system and increase perceptions of safety. Three sets of negatively reinforcing loops act to accelerate restraint use and clinician burnout from unintended negatively reinforcing cycles present in the system. Meanwhile, a set of positively reinforcing loops mitigate harm through cycles of mutual trust and control. These narratives are emergent relationships within the complex system produced from individual system components and their interactive relationships. As such, this initial model and these narratives act as hypothesis generation for future testing.

The emergence of the first two narratives highlight the fact that use of restraints may simultaneously cause both protective and harmful consequences for clinicians. Narrative 1 describes the balancing loop that provides the immediate sense of safety at the bedside, where use of restraints acutely decreases the number of agitated patients, which decreases likelihood of being assaulted, increasing perceptions of safety, and thus then decreasing subsequent use of restraint to stabilize the system. This narrative likely occurs quickly over minutes to hours within a shift during agitation encounters. At the same time, however, narrative 2 describes the process that drives a negatively reinforcing cycle of increasing task load and stress due to use of restraints that contributes to a perception of being unsafe over a more gradual period of time. Narrative 2’s influence of restraint use on a clinician’s task load may be more subtle and less immediate, as the urgent need to control symptoms during an agitation event may outweigh or overshadow the added work and stress involved with the placement of restraints and subsequent tasks associated with a restrained patient (e.g., clinical reassessment, documentation). However, this negatively reinforcing loop can gradually add strain to clinicians operating within the system that ultimately manifests as burnout over the course of months or years of exposure to placing restraints on patients. We found references to this tension between these two narratives from our previous qualitative data from staff members who describe a “patient care paradox”^23^ that creates a sense of moral injury and resulting stress when clinicians attempt to balance their own personal safety (narrative 1) and the desire to respect the safety and autonomy of patients (narrative 2).

Similarly, the negatively reinforcing loop of decreased support by colleagues and burnout in narrative 3 likely occurs over multiple episodes of agitation encounters amongst clinicians and develops gradually over time. Our prior work supports this narrative, as ED staff described lack of psychological safety and disparate goals regarding management decisions as key drivers for frustration and tension during agitation encounters.^14^ On the other hand, the positively reinforcing loop in narrative 5 appears to counteract these challenges through synergistic build-up of mutual trust, aligned perceptions of a physician’s control of the team, decreased physician stress, and resulting aligned perception of safety between nurses and physicians. Experts have called for better methods to support team-based care as means to reduce clinician burnout,^44^ and interprofessional care models show promise as potential methods to prevent long-term adverse effects on frontline clinicians at the highest risk of burnout and emotional exhaustion.^44^

Narrative 4 describes the negatively reinforcing loops of burnout and negative perceptions of patient intent that influence decisions on physical restraint use. Agitated patients often have substance use disorders, serious mental illnesses, and disadvantaged socioeconomic backgrounds, representing the most marginalized populations presenting to the ED.^45-47^ Unfortunately, these patients can be challenging to properly diagnose and treat due to difficulties in obtaining accurate histories and physical exams and establishing therapeutic rapport during decompensation of their underlying conditions. In addition, implicit bias and stigma against mental illness and substance use can further impede objective and patient-centered management decisions,^24,48,49^ and these clinician sentiments may heighten in the face of learned helplessness and emotional exhaustion from repeated exposure to workplace violence.^50^ Recent studies have shown that patients are able to perceive differences in bedside manner due to clinician burnout,^51^ and more work is needed to support the empathy and emotional bond necessary for de-escalation and behavioral techniques to be successful during agitation encounters.^52^

## LIMITATIONS

This study has some limitations that may affect generalizability. First, we included only the nursing and physician professions in the model. We acknowledge that many other professions experience burnout and play critical roles in the management of agitation, with key roles performed by patient care technicians and security officers during placement of physical restraints. In addition, we limited care provision in this model to within the ED only, excluding prehospital factors and transitions of care into the ED. Given the complexity that exists in the relationships between agitation and healthcare workplace violence, we chose to methodically start our process with contexts and professional roles that may exert the strongest influence in the model and be most proximal to decisions around use of restraints. Future work will include expansion of the model to encompass a broader system of factors that contribute to agitation management and may be amenable to interventions related to burnout and safety.

Some of the factors included in the model may lack consistent definitions in the literature or lack standardized measurement instruments (e.g., perception that the patient is problematic). Although these limitations may add barriers to incorporating currently available quantitative data in the model, future studies can elucidate new methods or tools to define and standardize these important factors identified in our work. We truncated some flows and simplified relationships between factors in the diagrams depicting our model for ease of visual interpretation and improved focus on the most pertinent aspects of relationships in the model. In addition, different sets of factors and flows may represent different time scales. For example, a clinical agitation encounter may occur over seconds to minutes, while mutual trust and burnout may change more gradually over months to years. To account for these considerations, we established significantly more detailed relationships, granular flows, and temporal factors for the analytic version of the model included in the Appendix to accurately incorporate quantitative data in future mathematical simulation studies. Finally, we incorporated expertise and previous data from two institutions only. We hope that our model offers a starting point to describe agitation and clinician burnout that incorporates a wider range of geopolitical and institutional experiences.

## CONCLUSIONS

Improving the management of agitated patients requires a balanced approach to clinician and patient safety.^15^ Using qualitative systems dynamics methods, we developed a new model illustrating the complex relationships between clinician experiences of assault, stress and burnout, and team interactions including mutual trust and how they impact decisions to restrain agitated patients. Consensus recommendations suggest minimizing the use of restraints in agitated patients.^3^ Yet, our model illustrates the importance of addressing clinician and system factors including workplace assault, burnout, stress, and team-based factors such as mutual support which each influence individual decisions to restrain an agitated patient. Our initial model serves as a first step in our SD modeling process. In future work, we will incorporate existing data, as well as prospective data collection, into a formal mathematical simulation of physical restraint use and clinical burnout over time in the system. We hope that our novel insights into the five clinical narratives identified in this current work will further support testing of potential interventions addressing both clinician burnout and reduction of restraint use.

## Data Availability

At the time of publication of any manuscripts that arise from this research, the de-identified data for that manuscript will be made available to share for scholarly activities. Qualitative data will be shared as a de-identified Dedoose dataset, and quantitative data will be shared as a de-identified CSV file. Sharing of the data will require a Data Use Agreement to be established between the requesting institution and Yale University. Data will be shared through secure file transfer.

## LIST OF ABBREVIATIONS

ED: emergency department
SD: system dynamics
WPV: workplace violence

## DECLARATIONS

### Ethics approval and consent to participate

The study received ethical approval from the Yale University human investigation committee as an exempt study (HIC# 2000028272, May 26, 2020). All methods were performed in accordance with the relevant guidelines and regulations.

### Consent for publication

Not applicable.

### Availability of data and materials

The datasets used and/or analyzed during the current study are available from the corresponding author on reasonable request.

### Author contributions

AHW, NSS, JMR, and RH conceived and designed the study; performed data extraction and synthesis; analyzed and interpreted the data. HRR performed external review and auditing of data collection and analysis. All authors drafted and contributed to critical revisions of the article. AHW takes responsibility for the paper as a whole.

## Acknowledgements

We would like to acknowledge Dr. Joseph Ross (Yale School of Medicine) for his contributions to the framing of this manuscript.

---

**Appendix 1.**
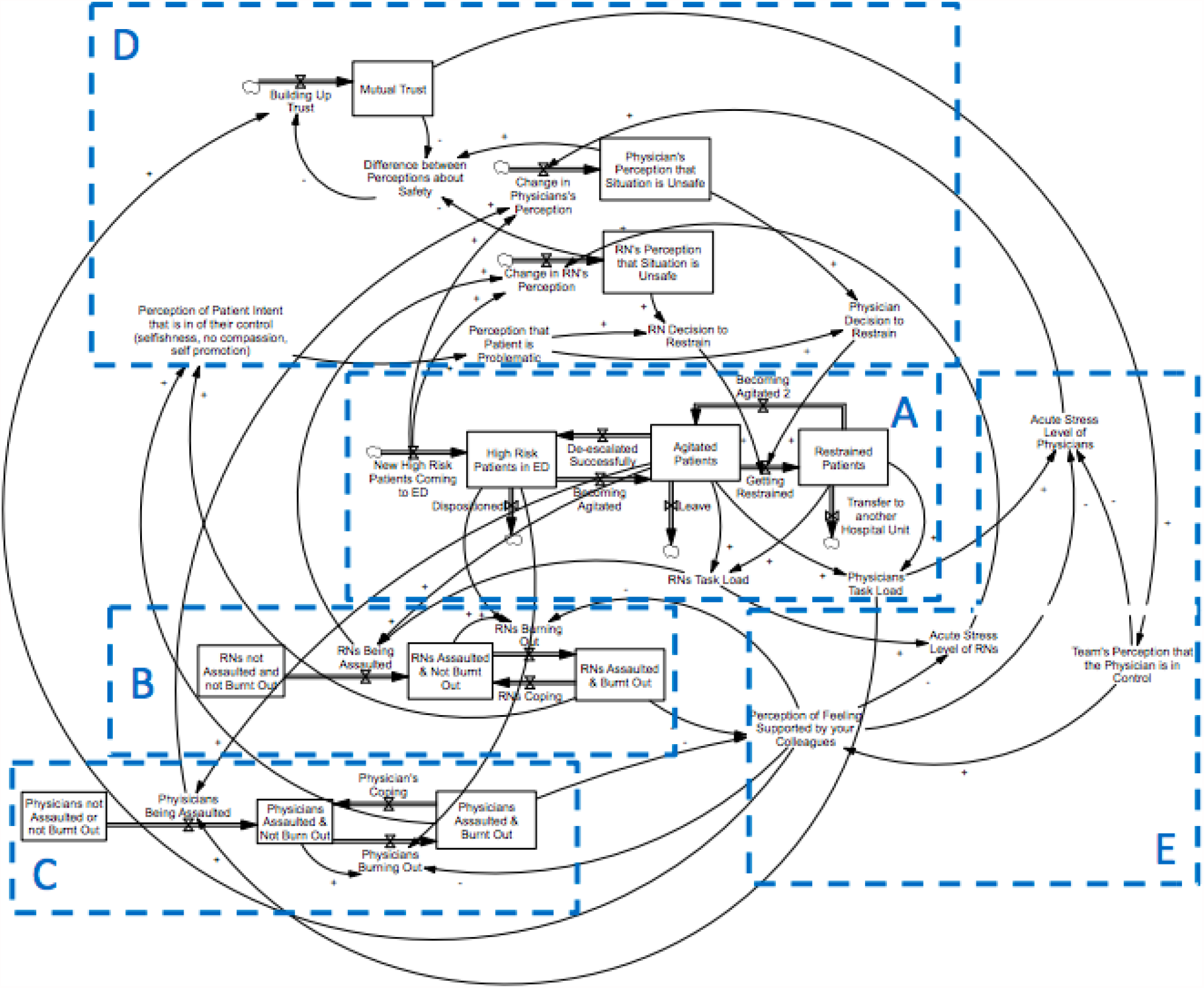
Detailed Qualitative System Dynamics Model for Agitation Management, Clinician Burnout, and Decisions for Physical Restraint Use. Identified sub-sections A) Patient Flow; B) RN (Registered Nurse) Flow; C) Physician Flow; D) Perceptions of Safety and Development of Trust, and E) Perceptions of Control.

**Appendix 2.**
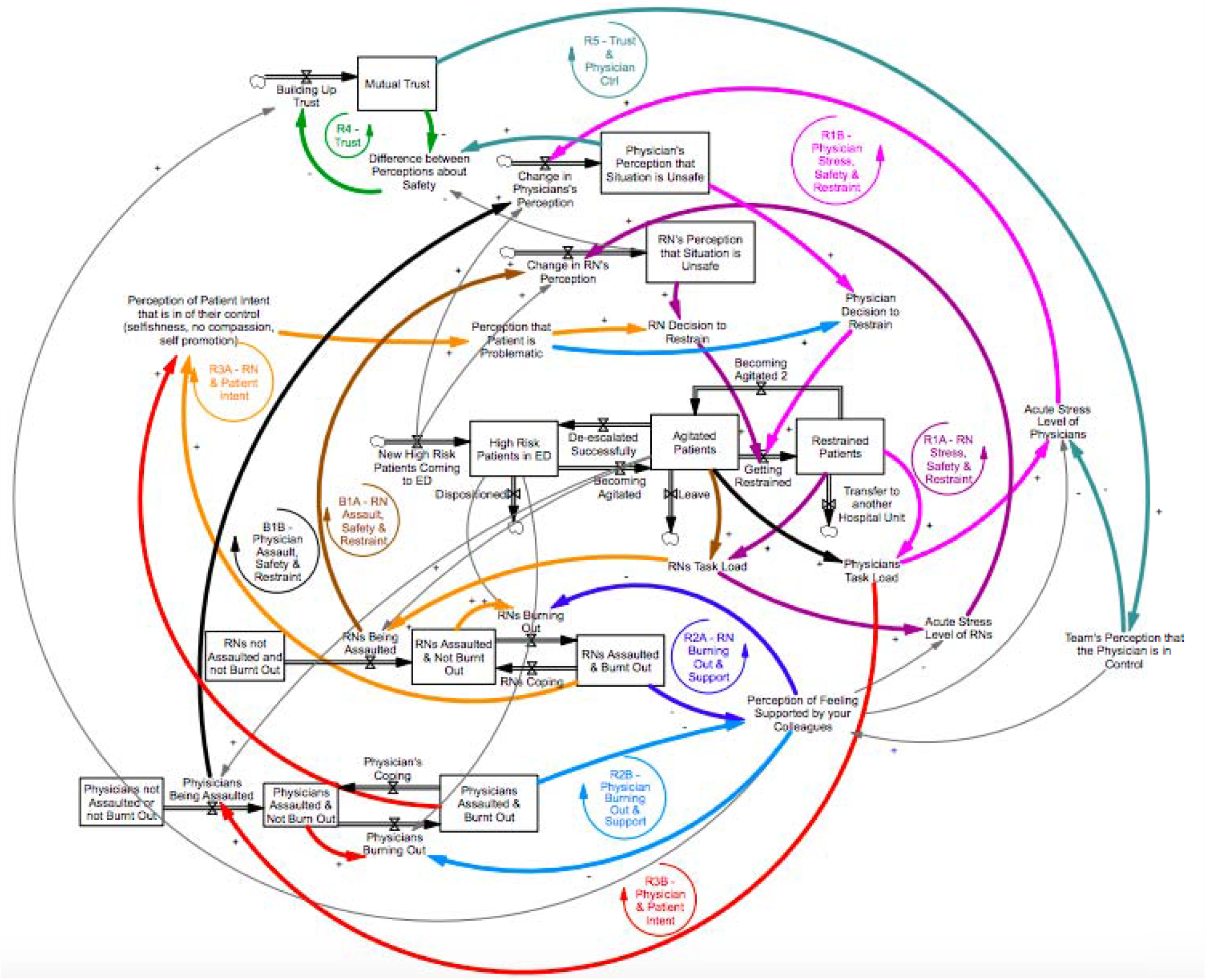
Highlighted Balancing and Reinforcing Loops within detailed qualitative model. The bolded black and brown loops (B1A/B1B) represent a pair of mirrored **balancing** loops, and other complementary colors represent four pairs of mirrored **reinforcing** loops. See Appendix 3 for details of each sets of loops.

**Appendix 3.**
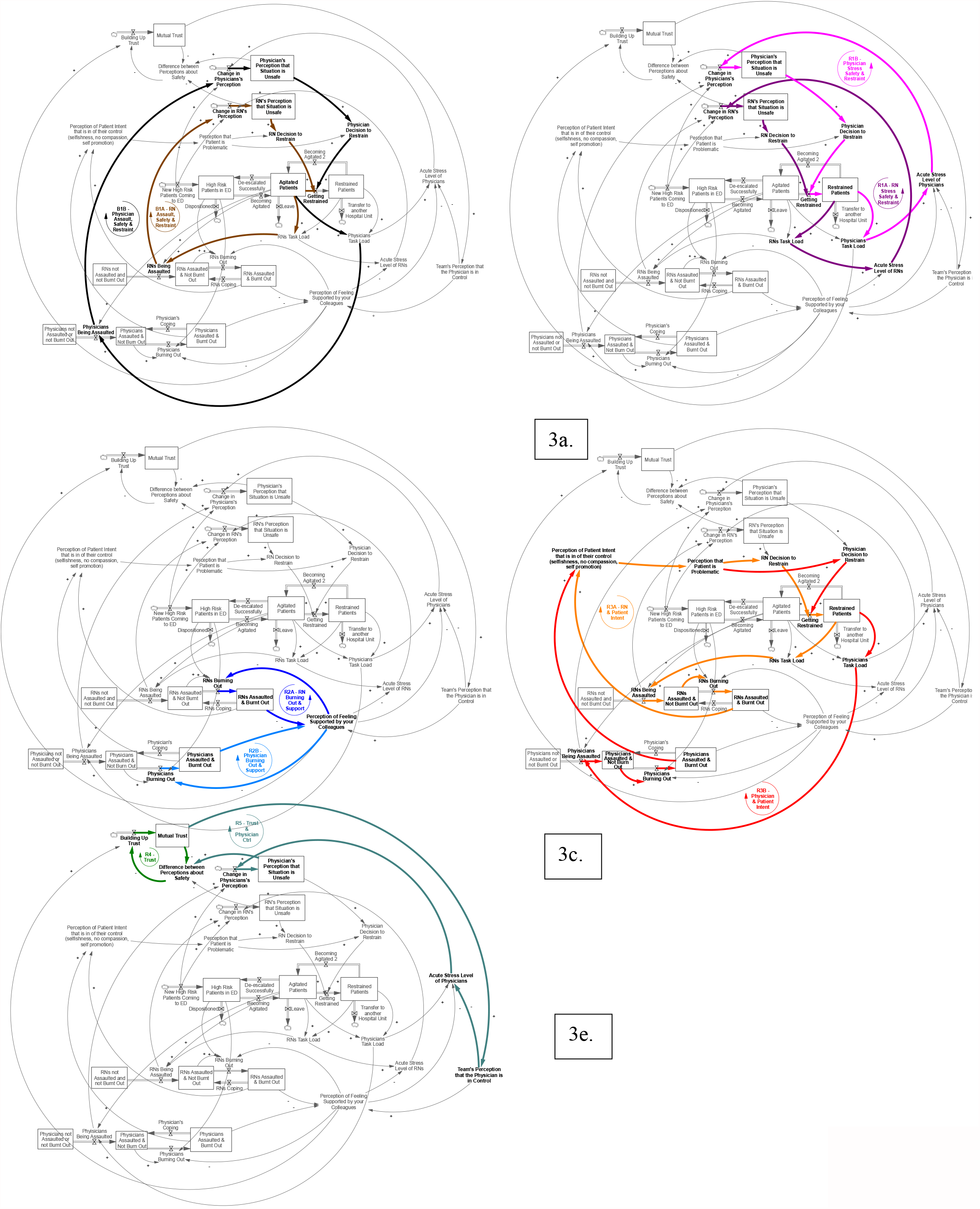
Individual Narratives within Detailed Model. 3a. Narrative 1—Use of Restraints, Assaults, and Perceptions of Safety (Balancing Loops B1A for Nurses and B2B for Physicians) 3b. Narrative 2—Clinician Stress, Safety, and Use of Restraints (Negatively Reinforcing Loops R1A for Nurses and R1B for Physicians) 3c. Narrative 3—Clinician Burnout and Support (Negatively Reinforcing Loops R2A for Nurses and R2B for Physicians) 3d. Narrative 4—Burnout & Perception of Patient Intent (Negatively Reinforcing Loops R3A for Nurses and R3B for Physicians) 3e. Narrative 5—Development of Trust and Control (Positively Reinforcing Loops R4 for Mutual Trust and R5 for Physician Control and Trust)

